# Homeostatic inflammation in the placenta is protective against adult cardiovascular and depressive outcomes

**DOI:** 10.1101/2023.02.20.23286171

**Authors:** Eamon Fitzgerald, Mo Jun Shen, Hannah Ee Juen Yong, Zihan Wang, Irina Pokhvisneva, Sachin Patel, Nicholas O’Toole, Shiao-Yng Chan, Yap Seng Chong, Helen Chen, Peter D Gluckman, Jerry Chan, Patrick Kia Ming Lee, Michael J Meaney

## Abstract

Pathological placental inflammation increases the risk for several adult disorders, but these mediators are also expressed under homeostatic conditions, where their contribution to adult health outcomes is unknown. Here we define an expression signature of homeostatic inflammation in the term placenta and use expression quantitative trait loci (eQTLs) to create a polygenic score (PGS) predictive of its expression. Using this PGS in the UK Biobank we carried out a phenome-wide association study, followed by mendelian randomization and identified protective, sex-dependent effects of the placental module on cardiovascular and depressive outcomes. Genes differentially regulated by intra-amniotic infection and preterm birth were also over-represented within the module. Our data support a model where disruption of placental homeostatic inflammation, following preterm birth or intra-amniotic infection, contributes to the increased risk of depression and cardiovascular disease observed in these individuals. Finally, we identify aspirin as a putative modulator of this homeostatic inflammatory signature.

## Introduction

Early studies, foundational to the developmental origins of health and disease (DOHaD) hypothesis, described an increased risk of both cardiovascular disease and depression in those born at low birth weight ^1–5^. As the site of maternal-fetal interface during pregnancy, the placenta is a critical regulator of cardiovascular effects as well as those of mental health ^6,7^. For instance, monozygotic twins that share a placenta are up to 6-fold more likely to be concordant for schizophrenia than monozygotic twins with separate placentae ^8^. Genetic studies have further reinforced the essential role of the placenta in shaping adult health outcomes ^9–12^, with the role of placental inflammation being particularly compelling ^13–16^.

A large body of literature in model systems supports a causal role for prenatal infection in driving adverse adult behavioural outcomes (reviewed by Estes and McAllister, 2016; Han *et al*., 2021), but the inflammatory mediators involved in the placental response to infection are also highly expressed under healthy conditions ^19,20^. Work in the developing brain with microglia illustrates the important homeostatic role of inflammatory mediators, which traditionally were thought to subserve functions solely related to infection ^21^. For instance, microglia are yolk sac derived macrophages and the placenta is home to its own yolk sac-derived macrophage, Hofbauer cells. Like microglia these cells have important homeostatic roles ^22,23^, but also display a robust response to infection ^22,24–26^. Little has been done to identify the role of these cells or gene expression patterns related to placental homeostatic inflammation. This is surprising as understanding homeostatic functions would also likely be informative for disease mechanisms.

In this study, we hypothesized homeostatic patterns of placental inflammation would shape adult health outcomes in offspring. To test this hypothesis, we performed a series of experiments using the Singapore-based, Growing Up in Singapore Towards healthy Outcomes (GUSTO) and UK Biobank cohorts. We first used RNA sequencing from 42 placental villous samples, obtained as part of the GUSTO study, and used weighted correlation network analysis (WGCNA ^27^) to identify a gene co-expression module associated with homeostatic inflammation. We discovered a module highly enriched in Hofbauer cells. We leveraged previously identified placental expression quantitative trait loci (eQTLs) ^28^ to generate a polygenic score (PGS; henceforth referred to as fetoplacental PGS), which specifically predicted expression of the genes comprising this module. To explore the functional relevance of this PGS we conducted a phenome-wide association study (pheWAS) in the UK Biobank and identified significant associations (false discovery rate; FDR<0.05) with 21 traits primarily within the cardiometabolic and mental health domains. We then used logistic regression and Mendelian randomization analyses to demonstrate protective sex-dependent effects on cardiovascular disease and depression related outcomes. Next, we demonstrated that our placental module was highly enriched for genes differentially regulated by intra-amniotic infection and preterm birth, and that these genes were among the most highly connected within the network. These data support a model by which loss or disruption of Hofbauer function as a consequence of preterm birth or intra-amniotic infection, respectively, contributes to the increased risk of depression and cardiovascular disease observed in these individuals ^29–33^. Finally using the Drug-Gene Interaction database (DGIdb), we identify aspirin as a promising candidate that may have therapeutic value when used prophylactically in populations at high risk of intra-uterine infection.

## Methods

### Cohorts

We used data from two population-based cohorts. The first was the Growing Up in Singapore Towards healthy Outcomes (GUSTO; Soh *et al*., 2014) cohort, a Singapore-based longitudinal birth cohort that recruited mothers at least 18-years of age from the two largest maternity units in Singapore. Ethical approval for GUSTO was granted by the relevant institutional boards (DSRB reference D/09/021 and CIRB reference 2009/280/D) and written informed consent was received from all participating mothers. The second data source was the UK Biobank, a large adult population-based study in the UK. The UK Biobank is guided by an ethics advisory committee and informed consent was received from all participants. Approval for the UK Biobank was obtained by the Northwest Multicentre Research Ethics Committee (REC reference 11/NW/0382), the National Information Governance Board for Health and Social Care and the Community Health Index Advisory Group. Access to data used in the current study was obtained under application #41975. In all analyses performed, sex was defined genetically. Samples sizes for specific analyses are described in the relevant results or supplementary tables.

### Placental sampling

Placenta samples (n=44; 2 subsequently excluded with hierarchical clustering) used in this study were derived from term births with tissue collected within 40 minutes of delivery. Exclusion criteria included antenatal smoking (confirmed with plasma cotinine Ng *et al*., 2019), maternal BMI greater than 30kg/m^2^, antenatal fasting glucose greater than 7 mmol/L or 2 hour oral glucose tolerance test result greater than 11.1 mmol/L, hypertensive disorders of pregnancy, birth prior to 37 weeks of gestation and a gestational age and sex-standardized birthweight percentile less than 10%. Placental biopsies were taken at random from 3 sites at the maternal-facing side, before removal of the maternal decidua to primarily retain the placental villous tissue for analysis. Sampling sites were then pooled and stored at −80C until further processing.

### RNA extraction and sequencing

Total RNA was extracted from samples using the phenol-chloroform method, followed by large RNA purification using the NucleoSpin miRNA kit (Machery-Nagel, Düren, Germany) as per manufacturer’s instructions. RNA concentrations were determined using a Nanodrop spectrophotometer (Thermo Fisher Scientific, Waltham, MA, USA) and RNA integrity number (RIN) was measured using the Agilent 4200 TapeStation System (Santa Clara, CA, USA). Sequencing libraries were prepared from samples with a RIN > 6 at Novogene AIT Genomics (Singapore). In brief, ribosomal RNA was depleted with the Illumina Ribo-Zero Magnetic Kit for Human/Mouse/Rat (San Diego, CA, USA). Library preparation was done using the NEB Next Ultra Directional RNA Library Prep Kit (New England Biolabs, Ipswich, MA, USA), before sequencing was carried out using the Illumina HiSeq platform with a minimum depth of 50 million paired-end 150bp reads. Sequencing quality was assessed using FastQC ^35^ and MultiQC ^36^. Reads were aligned to the human hg19 genome and gene level counts assembled with STAR Aligner ^37^. Samples had an average unique mapping rate of 93.5%.

### WGCNA

First raw gene counts were converted to RPKM (Reads Per Kilobase of transcript per Million mapped reads). Genes with missing information from more than 10% of samples were excluded. Agglomerative Hierarchical Clustering and Euclidean distance were used to assess outliers. After which 42 samples and a total of 31,097 genes remained. Gene co-expression networks were constructed by automatic block-wise network construction and the module detection process implemented in the WGCNA package ^27,38^. The soft thresholding power (β =14) for adjacency calculation was chosen based on approximate scale-free topology before co-expression network construction. The topological overlap matrix (TOM) was built by calculating gene adjacencies using biweight midcorrelation ^38^ with the chosen soft thresholding power, before signed weighted correlation networks were built. Modules were detected via hierarchical gene clustering on TOM-based dissimilarity and branch cutting using the top-down dynamic tree cut method ^39^. Eigengenes were calculated for each module and modules with high eigengene correlations (r > 0.85) were merged. Genes not incorporated to any module were assigned to the non-functional grey module. In total 28 functional modules were identified.

### Module preservation

*Z*_*summary*_ statistics were calculated for module preservation analysis by combing module density-based statistics and intra-modular connectivity-based statistics and separability of modules based on permutation test 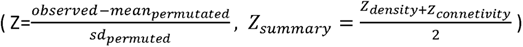. As per the recommended thresholds we defined *Z*_*summary*_ < 2 as no evidence of preservation, 2< *Z*_*summary*_ < 10 as weak to moderate evidence of preservation and *Z*_*summary*_ > 10 as strong evidence of preservation ^40^.

We tested our modules for preservation in an independent dataset, GSE148241 ^41^, which included 41 placenta samples (9 with early-onset severe preeclampsia and 32 healthy controls). Only the 32 healthy control samples were extracted for use in the analysis. We performed 100 permutations to reconstruct the networks using the same parameters to randomly permute module assignment in the test data.

### Gene ontology

Gene ontology enrichment for biological processes was carried out using the Gprofiler online interface with the default settings ^42^.

### Single cell and cell type enrichment

Cell type enrichment in datasets from Vento-Tormo *et al* ^19^ and Suryawanshi *et al* ^*20*^ was carried out using the PlacentaCellEnrich tool ^43^.

For single cell co-localization, normalized scRNA seq data from Vento-Tormo *et al* 2018, was downloaded using the Single Cell Browser ^44^ (https://placenta-decidua.cells.ucsc.edu). Data were then scaled and visualized with Seurat v3.2.3 as previously described ^45^. A module score was created using a previously described method ^46^.

### Transcription factor enrichment analysis

Transcription factor enrichment analysis was performed using the Top Rank method in the ChEA3 package using the online interface and default settings ^47^.

### Genotyping and PGS generation

Genotyping in the GUSTO cohort was performed with the Infinium OmniExpress Exome array. Quality control was done separately for each genetic ancestry. SNPs with a call rate < 95%, minor allele frequency < 5%, that were non-autosomal or that failed the Hardy-Weinberg equilibrium (p-value of 10^−6^) were removed from the analysis. Variants discordant from their respective subpopulation in the 1000 Genomes Project reference panel were removed (Chinese-EAS with a threshold of 0.20; Indian-SAS with a threshold of 0.20; Malay-EAS with a threshold of 0.30).

Samples were removed if they had ancestry or sex discrepancies, call rate < 99% or showed evidence of cryptic relatedness. Data were then pre-phased with SHAPEIT v2.837 with family trio information, and imputation carried out using the Sanger Imputation Service with the 1000G Phase 3 dataset as a reference, using the “with PBWT, no pre-phasing” (the Positional Burrows Wheeler Transform algorithm) pipeline. Imputed SNPs common to all genetic ancestries, which were bi-allelic, non-monomorphic and that had an INFO score > 0.8 were retained for downstream analysis.

UK Biobank genotyping and quality control processes are comprehensively described in Bycroft *et al* 2018 ^48^. Participants were excluded from the analysis if consent was withdrawn, genotyping data was unavailable, a genetic kinship to other participants > 0.044 identified, inconsistent genetic and reported sex, or if the subject was an outlier for heterozygosity. We then identified a single participant from each genetic kinship group (genetic relatedness < 0.025), based on their genomic relationship matrix (calculated using Genome-wide Complex Trait Analysis GCTA 1.93.2), which were returned to the analysis.

### PGS generation

PGS based on gene lists were generated as previously described ^49–51^. In brief, SNPs located on genes in a relevant list were identified using the biomaRt package ^52,53^. SNPs common with placental eQTLs identified by Peng *et al*, 2017 were retained. These eQTLs were subjected to linkage disequilibrium clumping (r^2^⍰<⍰0.2) in GUSTO and the UK Biobank, leaving only independent loci. For PGS calculation, the number of effect alleles at a particular locus was weighted based on the effect size on gene expression identified by Peng *et al*, before summation within individuals to generate the relevant PGS.

PGS for cardiovascular disease (CVD) and major depressive disorder (MDD; without the 23 and me sample) was performed using GWAS summary statistics from Nikpay *et al* 2015 and Howard *et al* 2019 using PRSice software v2.2.11.b ^54^ at 10 p-value thresholds (0.00000001, 0.0000001, 0.000001, 0.00001, 0.0001, 0.001, 0.01, 0.1, 0.2 and 0.5).

### Single sample gene set enrichment analysis (ssGSEA)

ssGSEA was implemented through the GSVA package as previously described ^55^. Scaled RPKM gene counts were used in the analysis. The effect of the PGS on ssGSEA score in the GUSTO RNA-seq samples was determined through multiple regression using sex and the first 3 genetic principal components as co-variates in the analysis.

### Cord blood analysis

Molecular characterization of cord blood from the GUSTO cohort (n=194-251) was conducted in duplicate using commercially available assays. Samples were randomized across plates and internal controls were used to estimate cross-plate variation. Assays with a coefficient of variation exceeding 20% across internal standards were excluded. Molecular profiles were analyzed using 1 of 3 methods: single molecule array (SIMOA), DropArray and enzyme-linked immunosorbent assay (ELISA). **Table 8** describes the individual assays. SIMOA measurements were made using the SIMOA HD-1 Analyzer (Quanterix). DropArray measurements were made using the FlexMAP3D bead-based multiplex system (Luminex). Normalization was carried out across plates using a median centring method. Data with readings outside of the assay limits as indicated by the manufacturer were discarded.

### UK Biobank analysis

All analyses were restricted to unrelated individuals with self-reported sex matching genetically identified sex. Individuals with high genetic missing rates or heterozygosity were also excluded from the analysis.

We used the PHESANT package to explore associations with the PGS using a pheWAS framework in the UK Biobank, as described previously ^56^. For instances where an outcome was measured multiple times, the variable with highest sample size was retained.

### Mendelian Randomization

Mendelian randomization was conducted as previously described using the TwoSampleMR package v0.5.6 ^57,58^. In all MR analyses placental eQTLs were used as the exposure. Outcome SNPs from MDD ^59^, coronary heart disease ^60^ and myocardial infarction ^60^ GWAS were obtained through the IEU GWAS database. The MDD summary statistics without 23 and me subjects were used in the analysis. Sex-specific GWAS summary statistics for the PHQ-9 in European subjects were downloaded from the Neale lab website ^61^. Further information on this data can be found in **Table 19**. Exposure and outcome data were harmonized to the same effect allele, with ambiguous SNPs removed from the analysis. We then carried out a fixed effects meta-analysis of the genetic instruments using the IVW (inverse variance weighted) method. These results were then confirmed using the more conservative weighted median method which assumes at least 50% of the instruments used are valid ^62^. Heterogeneity and horizontal pleiotropy were then assessed using several methods including the Cochran Q statistic, leave one out analysis, single SNP analysis and the MR egger intercept ^63–66^

### Drug interactions

Drug-gene interactions were interrogated using the online interface of the Drug-Gene Interaction Database v4.2.0 ^67^.

### Enrichment analysis

Enrichment for differentially expressed genes in the cyan module was done using a Fisher’s exact test as implemented with the GeneOverlap package ^68^.

### Network connectivity

To investigate the connectivity of the genes in a particular module we used ARACNE (Algorithm for the Reconstruction of Accurate Cellular Networks ^69^) to identify significant gene by gene associations within the modules based on their mutual information. For each ARACNE-derived unweighted network, the connectivity scores of the hub genes were computed.

### Statistical analysis

All analysis unless otherwise stated were carried out using R v4.1.1 ^70^ and Rstudio v1.4.1717 ^71^. Regression analyses were used to analyze the association between the fetoplacental PGS and cord cytokines (GUSTO), ssGSEA (GUSTO) and UK Biobank outcomes using lm() or glm() functions. Specific covariates for each analysis are described in the dedicated figure legends. The cord blood measurements were log transformed before inclusion in a linear regression analysis with the fetoplacental PGS as the predictor. A significance threshold of 0.05 was used throughout, with the Benjamini-Hochberg method (False Discovery Rate; FDR) used to correct for multiple comparisons.

## Results

### Identification of 28 gene expression modules in term placental villous samples

We used bulk RNA sequencing followed by WGCNA in 44 term placental villous samples obtained as part of the GUSTO study to identify gene expression patterns associated with placental homeostatic inflammation. We sequenced samples consisting of Chinese (26 samples; 59%), Malay (7 samples; 16%) and Indian (11 samples; 25%) self-defined ethnicities, with 24 of the 44 sequenced samples being female. Sample characteristics are shown in **Table 1**.

Following hierarchical clustering of the RNA sequencing data, 2 samples were removed as putative outliers (**Figure S1A**) and 42 samples were submitted to WGCNA. From 31,097 expressed genes we identified 28 gene expression module (**Figure 1A**), ranging in size from 72 (skyblue module) to 6794 genes (turquoise module). Unassigned genes were grouped into the grey module, with the constituents of each module comprehensively described in **Table 2**.

**Figure 1.**
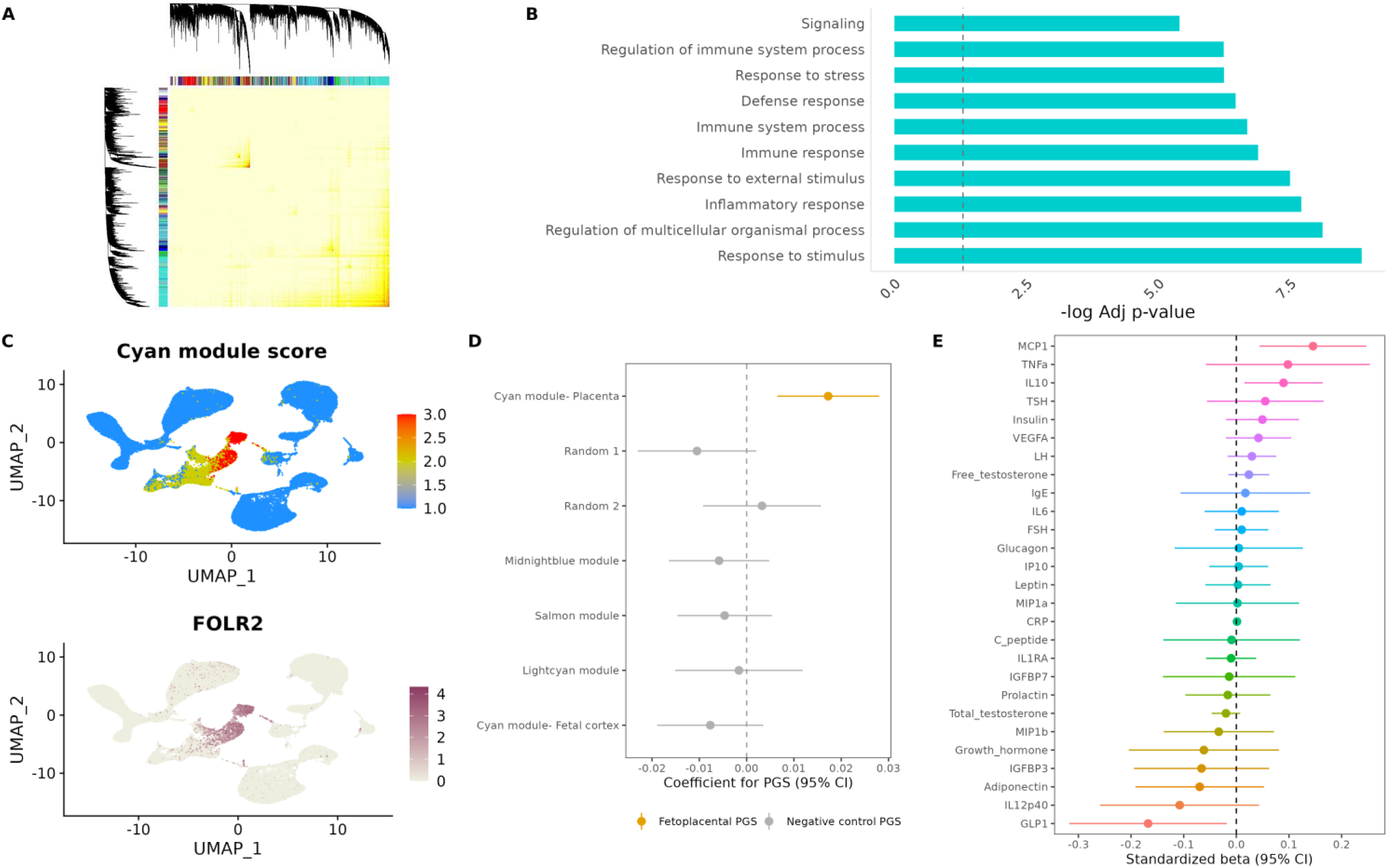
WGCNA of placental villous RNA sequencing data identifies a module of homeostatic inflammation that is highly enriched in Hofbauer cells. A) Heatmap of the topological overlap matrix, with corresponding dendrogram and module assignment (represented as colours). B) Gene ontology analysis of biological processes for the cyan module, ranked by −log of the adjusted p-value. Dashed line indicates FDR threshold for multiple comparisons. C) The top panel shows the cyan module score in scRNA-seq data from Vento-Tormo *et al*. The bottom panel shows expression of the Hofbauer cell marker *FOLR2*. D) Forest plot of the PGS coefficient from multiple regression analyses with 95% confidence intervals, estimating the effect of various PGS on single sample gene set enrichment analysis scores from the cyan module of the GUSTO RNA-seq dataset. The first 3 genetic principal components and sex were used as covariates in the analysis (n=42). E) Multiple regression analysis investigating the effect of the fetoplacental PGS on 27 molecules measured in cord blood. All outcomes were log transformed and scaled. The first 3 genetic principal components, sex and gestational age at birth were used as covariates in the analyses (n=194-251).

### Identification and characterization of a module related to homeostatic inflammation in the placental villous

The cyan module contained 486 genes (**Figure S1B**) and gene ontology analysis indicated a strong association with inflammation (**Figure 1B; Table 3**). The cyan module was also strongly preserved in an independent dataset ^41^ (**Figure S1C**). Several well characterized inflammation-related genes were present in the module including members of the tumor necrosis factor (*TNFRSF11A, TNFAIP8L2, TNFRSF21, TNFSF13, TNFSF9*) and interleukin (*IL2RA, IL12RB2, IL1RL1*) families, as well as several genes associated with the myeloid lineage (*AIF1, CSF1, CD33, CD163*). We found no evidence for a correlation between the cyan module expression and fetal sex (r= −0.037; p= 0.8).

Single-cell RNA sequencing data from the human placenta ^19,20^ showed cyan module genes were primarily expressed within Hofbauer cells (**Figure 1C and Figure S2A and 2B**), a fetal macrophage population localized to the placental villous ^72,73^. Transcription factor enrichment analysis also identified targets of transcription factors associated with macrophage identity, including *SPI1* and *MAFB*, as enriched within the cyan module (**Figure S2C**). Hofbauer cells are the predominant fetal immune cell in the placenta ^22^ with important homeostatic ^22,24–26^ and pathogenic roles ^22,23^. As such the cyan module presented an excellent candidate for further investigation.

We next used a placental eQTL resource from Peng *et al* ^28^ to identify SNPs contributing to variation in expression of cyan module genes (**Table 4**). In a comparison with the Gene-Tissue Expression (GTEx) v8 eQTL catalogue ^74^, the combination of these eQTLs associated with cyan module genes were highly specific to the placenta (**Figure S1D**). We then identified these variants in the children of the GUSTO cohort and weighted them based on their effect size from Peng *et al*, before summing them within individuals to create a fetoplacental PGS (as described previously ^49,50^). We reasoned that a PGS comprised of eQTLs would be representative of individual variation in cyan module expression. We validated this assumption using single sample gene set enrichment analysis in the GUSTO RNA-seq dataset (p=0.003, β=0.02, N=42; **Figure 1D**). A higher PGS score was associated with increased expression of cyan module genes. The cyan module PGS did not predict the expression of the other WGCNA modules (**Table 5**) and a PGS created in a similar fashion, using the WGCNA modules closest in size to the cyan module, had no effect on expression of cyan module genes (**Figure 1D; Table 6**). We provided additional controls for specificity by creating 2 PGSs based on randomly generated lists of 486 genes (same size of the cyan module) from our sequencing dataset and another using the cyan module but weighting it with eQTLs from fetal cortical tissue ^75^. None of these PGSs predicted expression of the cyan module (p=0.11, 0.61 and 0.19, respectively; **Figure 1D**). Taken together these findings suggest the fetoplacental PGS has considerable specificity for genes comprising the module.

Correlation analysis in the GUSTO cohort showed the fetoplacental PGS was not correlated with various perinatal factors (**Figure S1E**) including measures of maternal mental health, gestational age at birth, birth weight, offspring sex and socio-economic status.

Considering the importance of secreted factors during fetal development, we next investigated the association of the fetoplacental PGS with 27 cord blood molecules. These factors included cytokines (e.g. TNFα and IL6) and hormones (e.g. testosterone and insulin) with well-established prenatal effects. Using the fetoplacental PGS allowed us to expand our investigation to the entire sample of the GUSTO cohort with available genotype and cord blood data (n=194-251). Using the fetoplacental PGS as the predictor in a multiple regression analysis, the strongest effect was an association with monocyte chemoattractive protein 1 (MCP-1, also known as CCL-2; p=0.005, β=0.14; **Figure 1E; Table 7**). Interestingly, Hofbauer cells secrete high levels of MCP-1 ^22^, indicating our fetoplacental PGS may reflect Hofbauer cell function.

### Phenome-wide association study identifies adult cardiometabolic and mental health related outcomes are associated with the fetoplacental PGS

There is a paucity of studies that have evaluated placental contributions to adult outcomes of the offspring in large human datasets. We therefore created a PGS in the UK Biobank, using identical criteria to the previously described fetoplacental PGS in the GUSTO cohort. We used this PGS to perform a pheWAS with 1831 traits as outcomes of interests. Only unrelated individuals were used and outcomes were categorized to an appropriate regression family using the PHESANT package ^56^. We identified 21 significant associations (FDR p-value < 0.05; **Figure 2A; Table 9**) that were primarily localized to the cardiometabolic and mental health domains. All traits associated within the cardiometabolic domain had a positive direction of effect and all traits within the mental health domain had a negative direction of effect (**Table 9**). When samples were split by sex, no association passed the threshold for multiple comparisons, but both cardiometabolic and mental health traits approached the threshold for multiple comparisons in each sex (**Table 10 and 11**).

**Figure 2.**
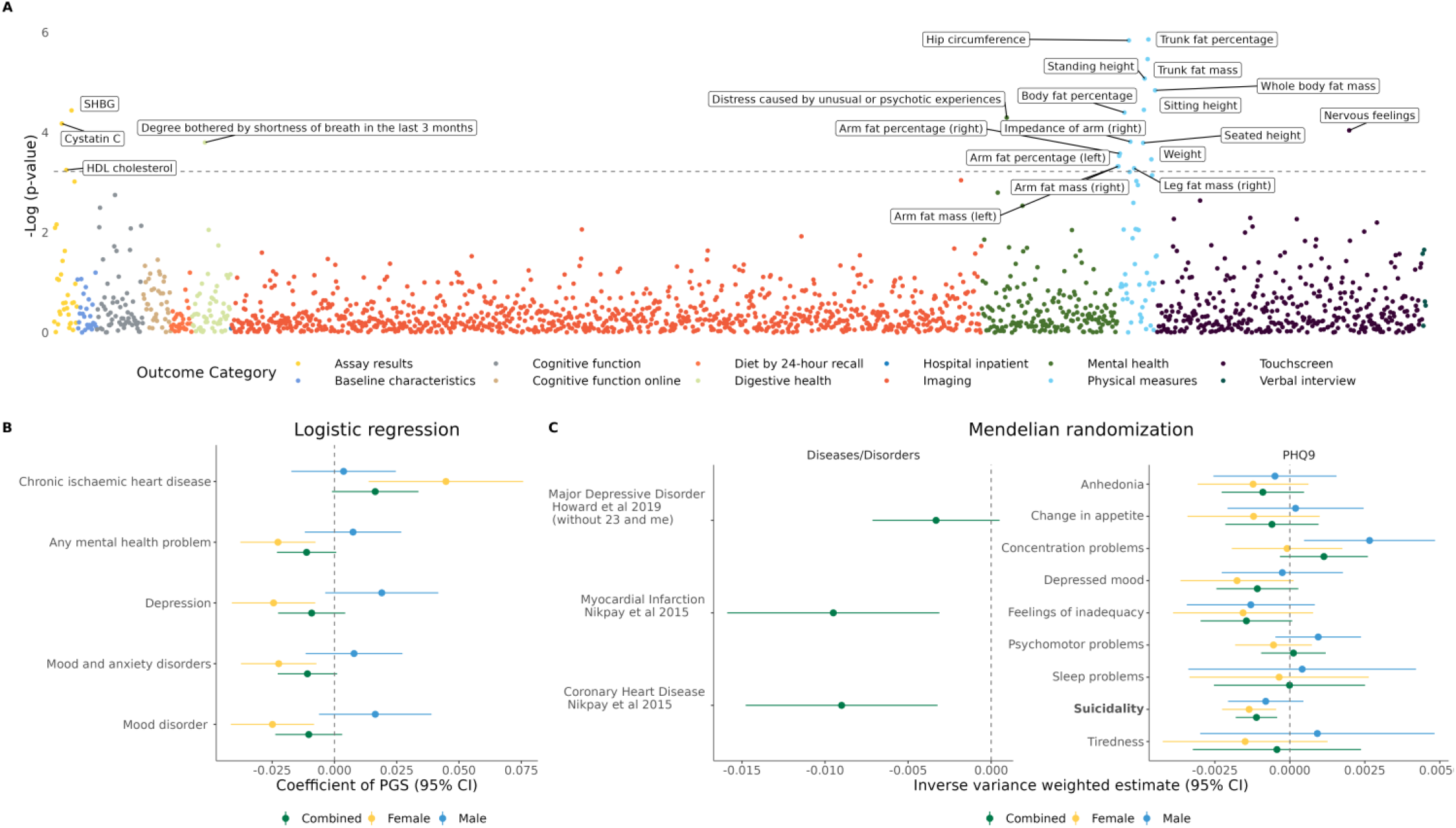
Phenome-wide association study and mendelian randomization analyses identify associations between the placental homeostatic inflammation module and adult outcomes. A) Manhattan plot showing the −log p-value corresponding to the effect of the fetoplacental PGS on 1831 outcomes in the full UK Biobank sample (males and females combined). Dashed line indicates the FDR threshold for multiple comparisons, all values exceeding this threshold are labelled. Points are colored as per their categorizations in the UK Biobank database. In each regression the first 10 genetic principal components, age, sex, genotype array and assessment center (categorical variable) are used as covariates. B) Logistic regression for the effect of the fetoplacental PGS on diagnoses in the UK Biobank. The x-axis represents coefficient with 95% confidence intervals, green represents male and female samples combined, yellow represents females only and blue represents males only. The first 40 genetic principal components, age, sex (only in combined sample), genotype array and assessment center (categorical variable) were used as covariates in the analyses. The 5 outcomes significantly associated with fetoplacental PGS after FDR correction in females are shown. C) Mendelian randomization using the inverse variance weighted method is used to analyze the effect of placental eQTLs on various outcomes. In both panels, green represents males and females combined, yellow represents females only and blue represents males only. The x-axis represents the IVW estimate with associated 95% confidence intervals. The first panel shows a significant effect for coronary heart disease and myocardial infarction (p= 0.002 and 0.004, respectively), with no significant association for MDD (p=0.09). The second panel illustrates the results from an analysis of responses to the PHQ-9, which are stratified by sex. Concentration problems in men (p=0.017) and suicidality in females (p=0.001) and the combined (p=0.003) were found to be significantly associated with placental eQTLs, but only suicidality in females (FDR=0.014) and the combined (FDR=0.028) samples retain significant association with placental eQTLs after correction for multiple comparisons. Suicidality is highlighted in bold font to denote an FDR p-value < 0.05.

Considering the striking enrichment of traits in the cardiometabolic and mental health domains, we next examined the association between the fetoplacental PGS and diagnoses within these domains. We considered 66 cardiometabolic and mental health diagnoses in the analysis and evaluated their association with the PGS in males, females and combined samples. We used logistic regression and identified 5 significant associations (FDR p-value<0.05), all of which were in females (**Figure 2B; Table 12-14**). Of these 5 associations, 4 pertained to mental health (“Any mental health problem”; FDR=0.049, β=−0.022, “Mood and anxiety disorders”; FDR=0.049, β=−0.022, “Mood disorder”; FDR=0.049, β=−0.024, and “Depression”; FDR=0.049, β=−0.024) and 1 to the cardiometabolic domain (“Chronic ischemic heart disease”; FDR=0.049, β=0.044). Depression was the most frequent diagnosis included under the 3 more broad mental health categorizations. We thus focused our subsequent analysis on depression and chronic ischemic heart disease. In line with our pheWAS results, the fetoplacental PGS had a protective effect for depression and acted to increase the risk for chronic ischemic heart disease. There was no correlation between our fetoplacental PGS and polygenic risk scores (PRS) for depression or cardiovascular disease (**Figure S3**).. Furthermore, the associations remained significant when a PRS for the relevant condition was used as a covariate in the analysis (**Figure S4**). The associations also remained significant when a diagnosis of depression or chronic ischemic heart disease was used as a covariate in the analysis of the other (**Figure S4**). These findings suggest our results are not the product of genetic architecture that overlaps with the relevant condition.

### Mendelian randomization identifies a protective effect of placental eQTLs for cardiovascular disease and suicidality in females

We next used Mendelian randomization as implemented in the TwosampleMR package ^57,58^. We used the eQTLs that composed the fetoplacental PGS as genetic instruments and the inverse variance weighted (IVW) method to estimate effects on both coronary heart disease and myocardial infarction ^60^. Contrary to our results using regression analyses, Mendelian randomization showed a protective effect on both outcomes (coronary heart disease, p=0.002, β=−0.009; myocardial infarction, p=0.003, β =−0.009; **Figure 2C; Tables 15**). The effect on myocardial infarction was confirmed with the weighted median method (p=0.01, β =−0.01), while coronary heart disease fell on the threshold for significance using this method (p=0.05, β =−0.008) (**Figure S5A and S6A; Table 16**). In supplementary analyses we found no evidence of instrument heterogeneity or horizontal pleiotropy using the Cochrane’s Q test, leave one out analysis, single SNP analysis and the MR egger intercept (**Tables 17 and 18; Figure S5B and S5C; Figure S6B and S6C**). This protective effect is in contrast to the increased risk we observed with regression analyses and underlines the importance of using orthogonal methods, such as mendelian randomization, that are more robust to environmental confounding.

Similar analysis for MDD also suggested a protective effect of the eQTLs, but was marginally outside the threshold for statistical significance using both the IVW (p=0.08, β =−0.003) and weighted median (p=0.08, β =−0.003) methods (**Figure 2C and S7; Table 15 and 16**). We reasoned this may either be due to sex-dependent effects, as observed in our logistic regression analysis, or to symptom specific effects. To address these questions we ran Mendelian randomization analyses using sex-specific GWAS summary statistics from the patient health questionnaire 9 (PHQ-9), answered as part of the UK Biobank ^61,76^. The PHQ-9 is a self-report questionnaire based on the 9 DSM criteria for a diagnosis of depression (**Table 19**). In this analysis, we identified a robust protective effect for suicidality in females (p=0.003, FDR=0.02, β =−0.001; **Figure 2C; Table 15**). This finding was confirmed with the weighted median method (p=0.04, β =−0.001; **Table 16**), with no evidence of heterogeneity or horizontal pleiotropy (**Figure S8; Table 17 and 18**). We also identified a significant effect in the full sample for suicidality using the IVW method, but this could not be confirmed by the weighted median method (**Figure S9; Table 17 and 18**).

These results demonstrate our expression signature of homeostatic inflammation in the placenta is protective against adult cardiovascular disease in a male/female combined sample and against suicidal ideation in females. These results are in line with a large body of literature describing a positive correlation between cardiovascular and depressive risk ^77,78^.

### Cyan module is highly enriched for genes differentially regulated by intra-uterine infection and preterm birth

Interestingly, previous work in populations exposed to instances of pathogenic inflammation, such as preterm birth or general prenatal infection, have shown an *increased* risk for both cardiovascular disease and depressive outcomes ^29–33^. This is in contrast to our findings, where we see *protective* effects of homeostatic inflammation on these outcomes. A parsimonious explanation for this is that pathological prenatal exposures may partly confer risk for adult health outcomes by disrupting the homeostatic function of inflammation related gene expression in the placenta.

Therefore, we next asked whether placental genes differentially regulated by preterm birth, infection or other exposures were enriched within the cyan module. We mined the published literature for studies that performed differential expression analysis in the human placenta following various exposures ^79–85^ and extracted differentially expressed genes. We found a very strong enrichment of genes differentially expressed in response to intra-amniotic infection (154 genes; p=5.2e-15) and preterm birth (15 genes; p=2.6e-05) in the cyan module (**Figure 3A; Table 20**). We then used ARACNE ^69^ to measure the connectivity of all genes within the cyan module and estimated the degree of connectivity in genes differentially regulated by intra-amniotic infection or preterm birth. This approach showed these genes were critical regulators of cyan module integrity and suggests severe disruption of the cyan module occurs with intra-amniotic infection (**Figure 3C; Table 21**).

**Figure 3.**
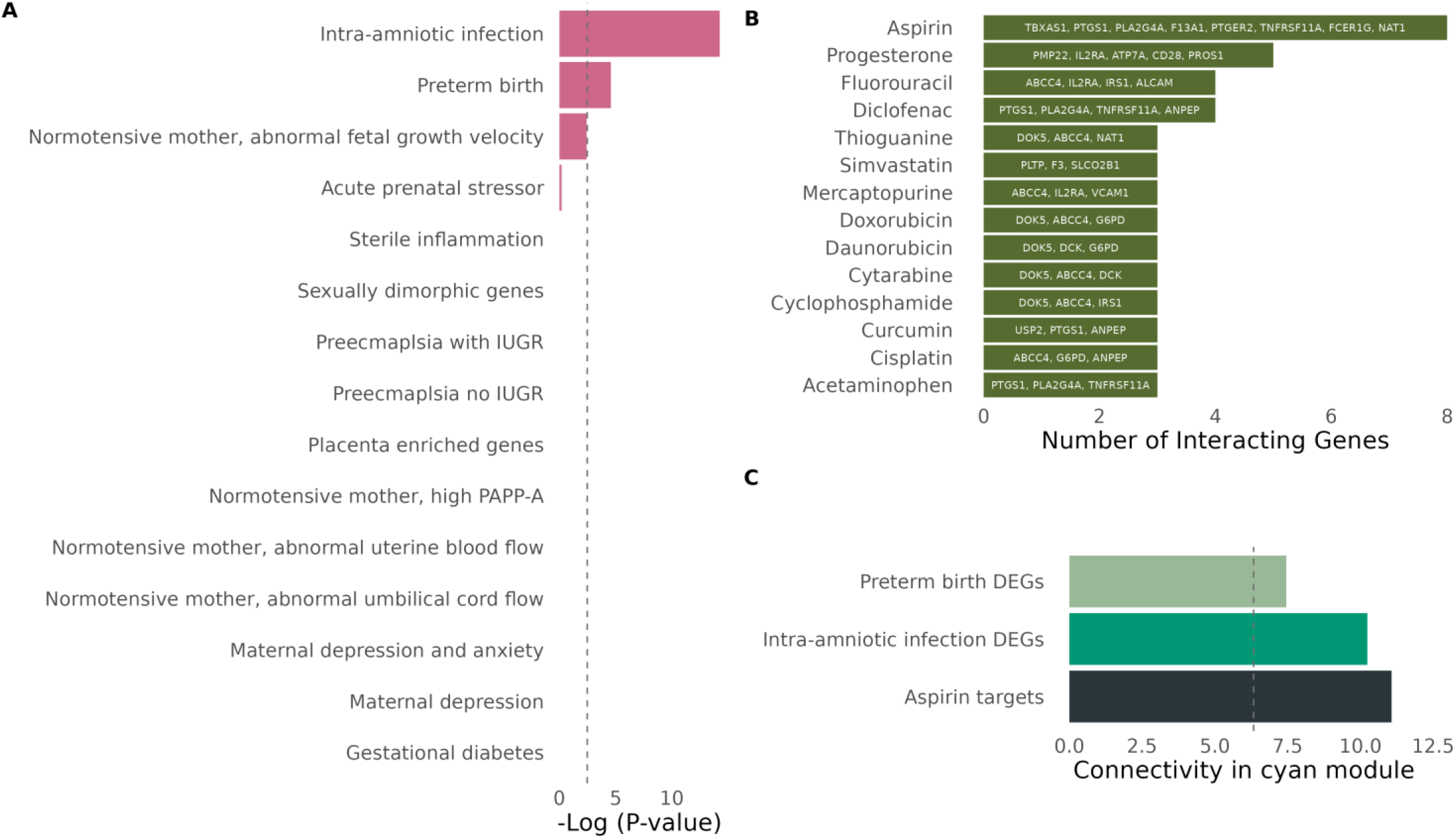
The placental homeostatic inflammation module is highly enriched for genes differentially regulated by intra-amniotic infection and preterm birth. A) Enrichment analysis for genes differentially expressed in the placenta under various conditions. Dashed line indicates threshold for multiple comparisons. B) Drugs annotated in the Drug-Gene Interaction Database which target members of the cyan module. Drug names are on the y-axis with the names of genes they target in the cyan module labelled within their respective column. C) Average connectivity score of genes differentially regulated by intra-amniotic infection, preterm birth or those found to be targets of Aspirin. Dashed line indicates the mean connectivity of all cyan module genes.

We finally asked if any drugs are to known to target genes of the cyan module. We used DGIdb, which compiles gene expression effects of drugs. This unbiased analysis identified the anti-inflammatory drug, aspirin, as the drug with the most targets within the cyan module (**Figure 3B**). Characterizing the connectivity of these genes demonstrated their essential importance to the module (**Figure 3C; Table 21**) and the potential for aspirin to modulate cyan module integrity.

Our data advocate for a mechanism by which intra-uterine infection and preterm birth confer risk for cardiovascular and depressive outcomes, at least partially, through disruption or loss of the homeostatic functions of the cyan module and the cell type within which it is primarily expressed, Hofbauer cells.

## Discussion

We used a multi-modal approach integrating transcriptomics, genetics and adult health outcomes across multiple cohorts and genetic ancestries to assess the effect of homeostatic placental inflammation on adult offspring outcomes. We used distinct complementary methods and several supplementary analyses to identify a novel protective effect of a placental gene co-expression module, principally expressed in Hofbauer cells, on adult depressive and cardiovascular outcomes. Our results suggest disruption or loss of the homeostatic functions served by Hofbauer cells may contribute to the increased risk of adult cardiovascular and depressive outcomes in individuals exposed to intra-uterine infection or born preterm.

Intra-uterine infection is a primary precipitant of preterm birth ^86^, which in turn is associated with a marked increase in risk for adult depression and cardiovascular disease ^29–31^. Loss of the placenta and therefore Hofbauer cell function, is an inevitable consequence of preterm birth. A model under which this premature loss of Hofbauer cell function contributes to adverse adult health outcomes in offspring is plausible. Indeed similar mechanisms have been described for several placental mechanisms, with particularly strong evidence for endocrine functions ^87,88^. Our cord blood analysis found the strongest association between the fetoplacental PGS and MCP-1, suggesting that it (or indeed other unmeasured secreted molecules) could act to stimulate fetal development. Premature cessation of this endocrine signalling may then contribute to adverse adult health outcomes in offspring. Hofbauer cells are understudied and future comprehensive characterizations of their function throughout pregnancy would likely further inform these mechanisms and may even point to novel therapeutic or preventive strategies in preterm infants.

Intra-uterine infection can also occur in pregnancies carried to term, but adult outcomes of this population is less studied. Population studies that do not discriminate between maternally confined and intra-amniotic infections have observed an increased risk for both depression and cardiovascular disease in offspring ^32,33^, but future work stratifying by infection type will be critical. In our study, the transcriptional response to intra-uterine infection was highly enriched within our Hofbauer gene expression module. These genes in turn were central components of the network, suggesting severe disruption in instances of intra-uterine infection. Histological studies have indeed seen changes in Hofbauer cell distribution with intra-uterine infection ^89^. Hofbauer cells also express physiologically functional Toll Like Receptors, suggesting an active role in the response to intra-amniotic infection ^22^. Previous studies have even observed direct infection of Hofbauer cells by HIV, Zika and SARS-CoV-2 viruses ^90–92^. Together, this suggests a novel clinical approach to reducing risk of adverse offspring health outcomes following intra-amniotic infection may be to target the cyan module and Hofbauer cells.

Of the drugs annotated in the DGIdb, the anti-inflammatory drug, aspirin, had the most targets in the cyan module. Even though these genes represented only a minority of the module’s membership the genes showed a very high degree of connectivity, indicating their potential to affect network integrity. Aspirin is an attractive candidate considering its breadth of use and volume of available data. In fact, aspirin is already recommended for use in pregnant women at high risk of preeclampsia ^93,94^, and estimates suggest it is already used by up to 38.8% of this population in the United States ^95^. It is unknown if aspirin therapeutically acts through Hofbauer cells in the context of preeclampsia, but a previously characterized function of Hofbauer cells is in angiogenesis. Disruptions in placental angiogenesis have also been observed with prenatal infection (reviewed by Weckman *et al*., 2019) and MCP-1, which we found to have the strongest association with the fetoplacental PGS in cord blood, is also an angiogenic regulator ^97,98^. A credible hypothesis based on our results is that intra-amniotic infection elicits a response from Hofbauer cells, which perturbs placental angiogenesis with long-term consequences for the fetus. The effects of aspirin on the cyan module, Hofbauer cell function, angiogenesis and ultimately whether it is beneficial for pregnancies at risk of intra-uterine infection, are key future questions.

We found several instances of sex-dependent effects in our study. This is not surprising, as sex differences in the prevalence of depression ^99,100^ and cardiovascular disease ^101^ have been widely reported. These conditions are also highly comorbid and genetically correlated ^77,78,102^. Our results provide evidence that sexually dimorphic regulation of risk for these outcomes starts during early development. We did not identify any correlation of cyan module expression with fetal sex and previous studies in other tissues have not observed notable sex-biases in eQTLs ^103^. Therefore, we have no evidence to suggest sex biases were present in our analyses at either the gene expression level or eQTL level. Processes downstream of the cyan module are then likely responsible for these sex-differences, but future work is required to establish the nature of these processes.

## Limitations

Our study has limitations that must be considered. First, our analysis was limited by the availability of placental functional genomic resources. Future studies with increased power for placental eQTL discovery are essential to increase the discovery power of studies such as ours. Second, due to the nature of many large GWASes, summary statistics were often only available for males and females combined. As GWAS sample size increases, we anticipate the wider availability of sex-specific summary statistics will further expand the study of sex differences into powerful methods like Mendelian randomization. Finally, we confined our RNA sequencing samples to term births to avoid pathological artifacts associated with preterm birth. Characterizing the developmental expression trajectories of homeostatic inflammation patterns in the placenta will be an important future step.

## Conclusions

In conclusion, we identified a gene expression module related to homeostatic inflammation in the placenta that was highly enriched in Hofbauer cells, and created a PGS that specifically predicted expression of this module. We used this PGS to identify associations with traits in the cardiometabolic and mental health domains of the UK Biobank. Follow-up analyses using logistic regression and Mendelian randomization demonstrated genetically-inferred cyan module expression was protective for cardiovascular and depressive outcomes. We finally showed that genes differentially regulated by both intra-amniotic infection and preterm birth were highly enriched in the cyan module. These findings suggest that loss of Hofbauer cell function with preterm birth or its disruption with intra-amniotic infection may contribute to the increased risk of cardiovascular disease and depression in these offspring.

## Data Availability

Access to data from the GUSTO and UK Biobank are dependent on approved application to the respective data access committees.

## Author contributions

EF planned the study, performed the analysis, interpreted the data and wrote the initial draft of the manuscript, with editing from MJM who provided funding. HC provided feedback on the manuscript. MJS performed WGCNA. HEJY and PL coordinated and prepared placental samples for RNA-seq. SYC oversaw the sample selection for RNA-seq. Placentas were collected in accordance with protocols developed by JC. RNA-seq data were analyzed by EF, NOT and MJM. Calculation of PGS was performed by ZW, SP and IP. YSC, PDG and MJM designed the GUSTO cohort study and obtained funding. All authors approved the final version of the manuscript.

## Data availability

Access to data from the GUSTO and UK Biobank are dependent on approved application to the respective data access committees. All other data generated in this study are provided in the supplementary material.

## Code availability

Code for these analyses was in line with vignettes for all packages mentioned in the methods. Code to run the pheWAS analysis can be found at https://github.com/MRCIEU/PHESANT. Code to run the Mendelian randomization analysis can be found at https://mrcieu.github.io/TwoSampleMR/.

## Acknowledgments

We would like to acknowledge both the GUSTO and UK Biobank investigators, staff, researchers and in particular the participants. This research has been conducted using the UK Biobank Resource under Application Number 41975.

## Figure captions

Supplementary Figure 1

**WGCNA and cyan module PGS**. A) Outlier removal with hierarchical clustering of placental villous samples prior to WGCNA. B) Cyan module network with nodes representing genes and color representing connectivity score. C) Preservation analysis of WGCNA modules in Yang *et al*. Above the dashed green line indicates strong evidence of preservation. D) Comparison of eQTLs used to generate the fetoplacental PGS (y-axis) in all GTEx v8 tissues (along x-axis; last value is the placenta eQTLs used in this study for comparison purposes). If an eQTL was identified in that tissue it is colored with respect to its normalized effect size (NES) as indicated in the GTEx catalogue. E) Correlation plot of the cyan module PGS and various perinatal factors. Empty box indicates no significant correlation at an uncorrected p-value threshold of 0.05. Correlations with a p-value <0.05 have a color and size proportional to their r.

Supplementary Figure 2

**Cell type enrichment and transcription factor enrichment of the cyan module**. A) Enrichment of the cyan module in scRNA-seq data from the Vento-Tormo **et al** (A) and Suryawanshi **et al** (B) both show strong enrichment in Hofbauer cells. Note these studies also included maternal macrophages, which are unlikely to be present in our dataset. C) Transcripton factor enrichment analysis of the cyan module using ChEA3 and the Top Rank analysis.

Supplementary Figure 3

**Correlation of fetoplacental PGS with major depression disorder (MDD) PRS and cardiovascular disorder (CVD) PRS**. Empty box indicates no correlation at an uncorrected p-value threshold of 0.05. Correlations with a p-value<0.05 have a color and size proportional to their r.

Supplementary Figure 4

**Logistic regression for female significant multiple regression diagnoses associations in the UK Biobank using diagnosis and PRS covariates**. Across all panels the x-axis represents the coefficient with 95% confidence intervals, yellow represents females only. The first 40 genetic principal components, age, genotype chip and assessment center are used as covariates in all analyses. The first panel displays the results of a logistic regression for depression in females when the covariates on the y-axis were separately added to the model. The second panel displays the results of a logisitic regression for Chronic ischaemic heart disease in females when the covariates on the y-axis were separately added to the model.

Supplementary Figure 5

**Scatterplot, single SNP and leave one out analysis for myocardial infarction**. A) Scatterplot of SNPs with their effect size for cyan module gene expression (x-axis) and myocardial infarction (y-axis). Dark blue line uses the weighted median method and the light blue line uses the IVW method. B) Single SNP analysis for individual SNPs (in black using the Wald ratio) and combined analysis using the IVW method (red). C) IVW results of the analysis when each SNP is sequentially removed from the analysis. Removed SNP indicated on the y-axis, combined IVW for all SNPs is in red. IVW; Inverse Variance Weighted.

Supplementary Figure 6

**Scatterplot, single SNP and leave one out analysis for coronary heart disease**. A) Scatterplot of SNPs with their effect size for cyan module gene expression (x-axis) and coronary heart disease (y-axis). Dark blue line uses the weighted median method and the light blue line uses the IVW method. B) Single SNP analysis for individual SNPs (in black using the Wald ratio) and combined analysis using the IVW method (red). C) IVW results of the analysis when each SNP is sequentially removed from the analysis. Removed SNP indicated on the y-axis, combined IVW for all SNPs is in red. IVW; Inverse Variance Weighted.

Supplementary Figure 7

**Scatterplot, single SNP and leave one out analysis for major depressive disorder**. A) Scatterplot of SNPs with their effect size for cyan module gene expression (x-axis) and major depressive disorder (y-axis). Dark blue line uses the weighted median method and the light blue line uses the IVW method. B) Single SNP analysis for individual SNPs (in black using the Wald ratio) and combined analysis using the IVW method (red). C) IVW results of the analysis when each SNP is sequentially removed from the analysis. Removed SNP indicated on the y-axis, combined IVW for all SNPs is in red. IVW; Inverse Variance Weighted.

Supplementary Figure 8

**Scatterplot, single SNP and leave one out analysis for suicidality (females only)**. A) Scatterplot of SNPs with their effect size for cyan module gene expression (x-axis) and suicidality (females only; y-axis). Dark blue line uses the weighted median method and the light blue line uses the IVW method. B) Single SNP analysis for individual SNPs (in black using the Wald ratio) and combined analysis using the IVW method (red). C) IVW results of the analysis when each SNP is sequentially removed from the analysis. Removed SNP indicated on the y-axis, combined IVW for all SNPs is in red. IVW; Inverse Variance Weighted.

Supplementary Figure 9

**Scatterplot, single SNP and leave one out analysis for suicidality (males and females combined)**. A) Scatterplot of SNPs with their effect size for cyan module gene expression (x-axis) and suicidality (males and females combines; y-axis). Dark blue line uses the weighted median method and the light blue line uses the IVW method. B) Single SNP analysis for individual SNPs (in black using the Wald ratio) and combined analysis using the IVW method (red). C) IVW results of the analysis when each SNP is sequentially removed from the analysis. Removed SNP indicated on the y-axis, combined IVW for all SNPs is in red. IVW; Inverse Variance Weighted.

