## Supplementary figures for "Homeostatic inflammation in the placenta is protective against adult cardiovascular and depressive outcomes"

Supp Figure 1

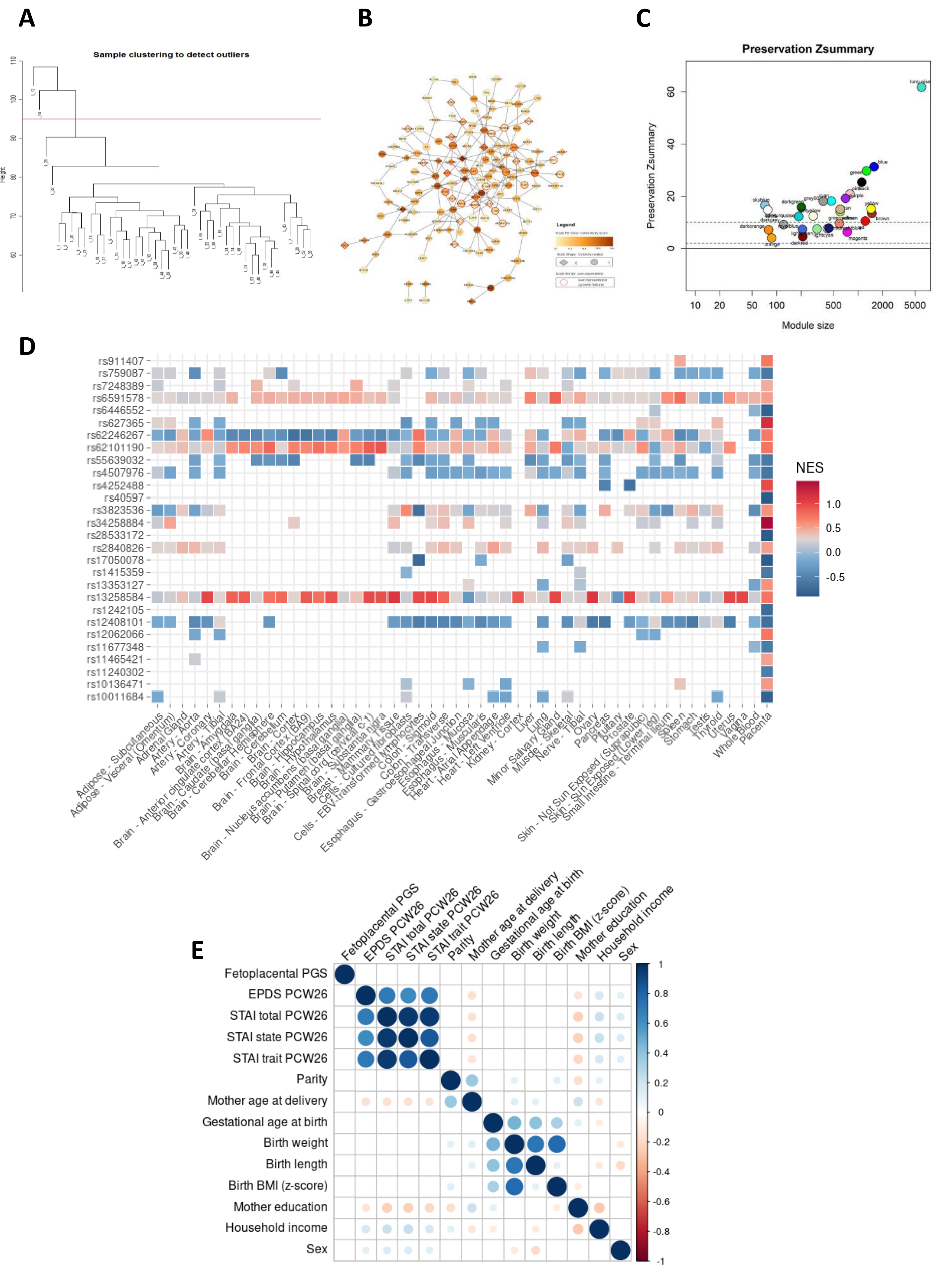

Supp Figure 2

A

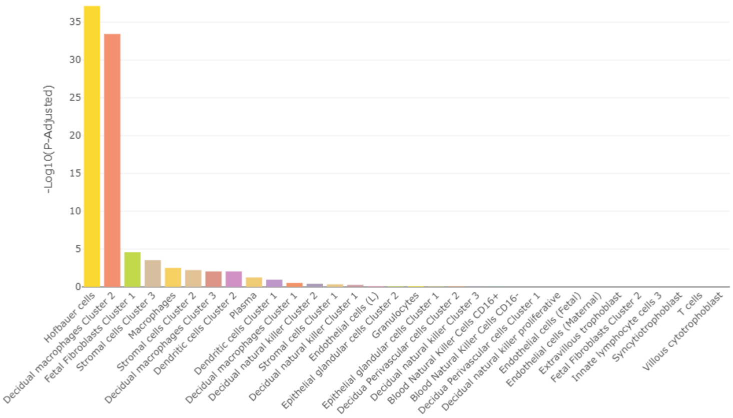


B

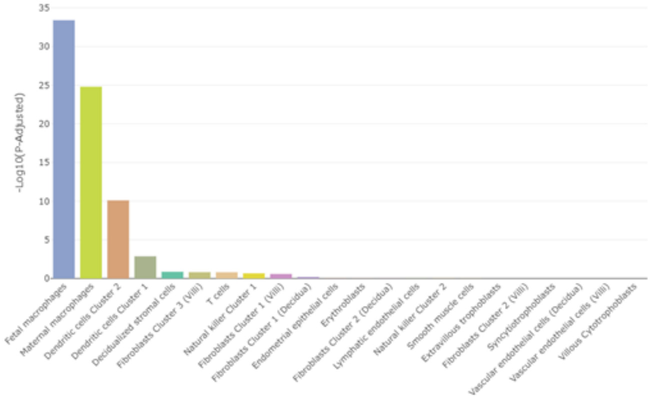


C

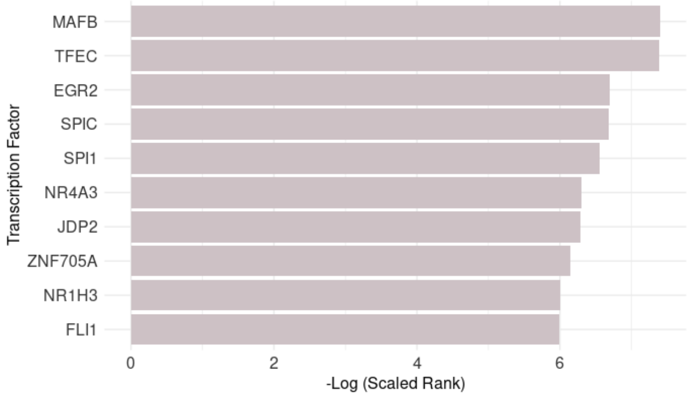

Supp Figure 3

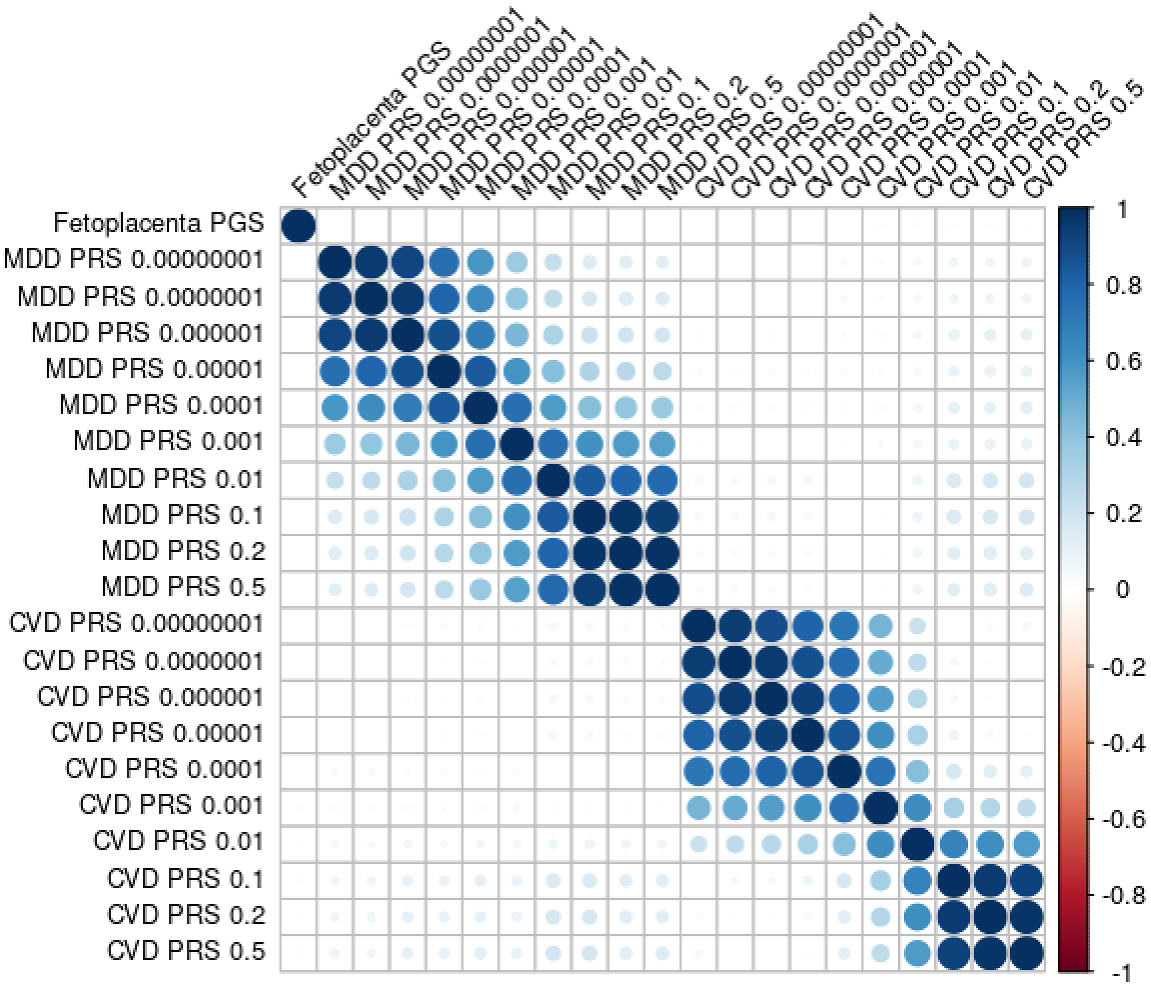

### Supp Figure 4

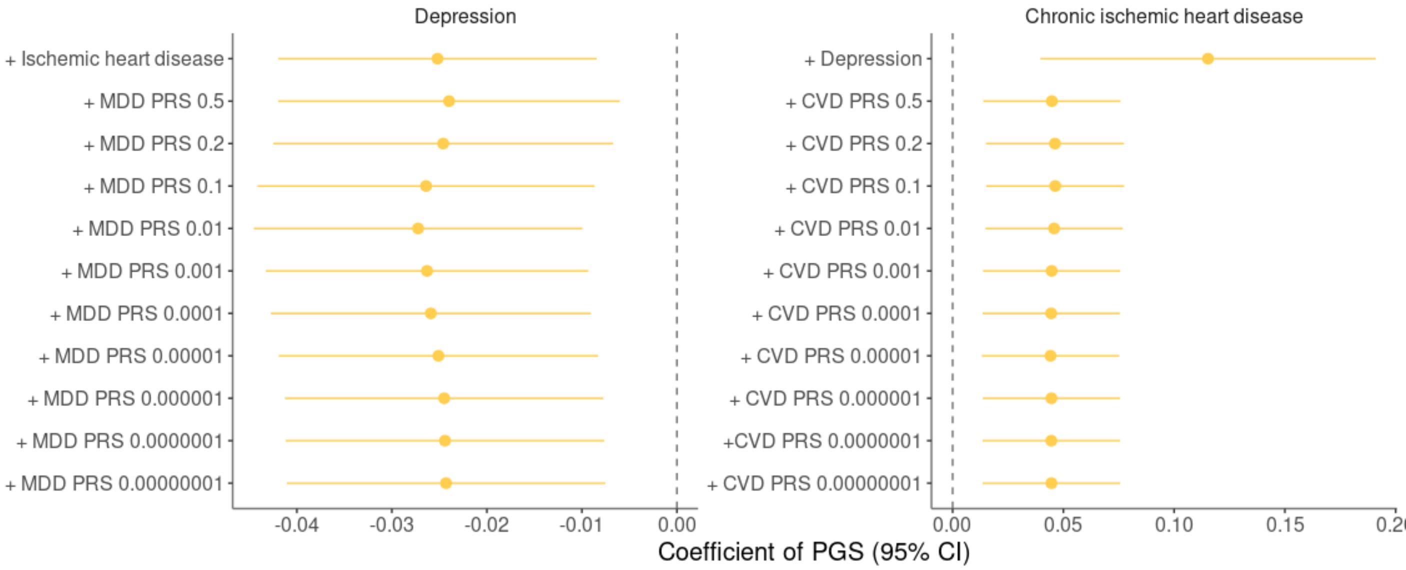

Supp Figure 5

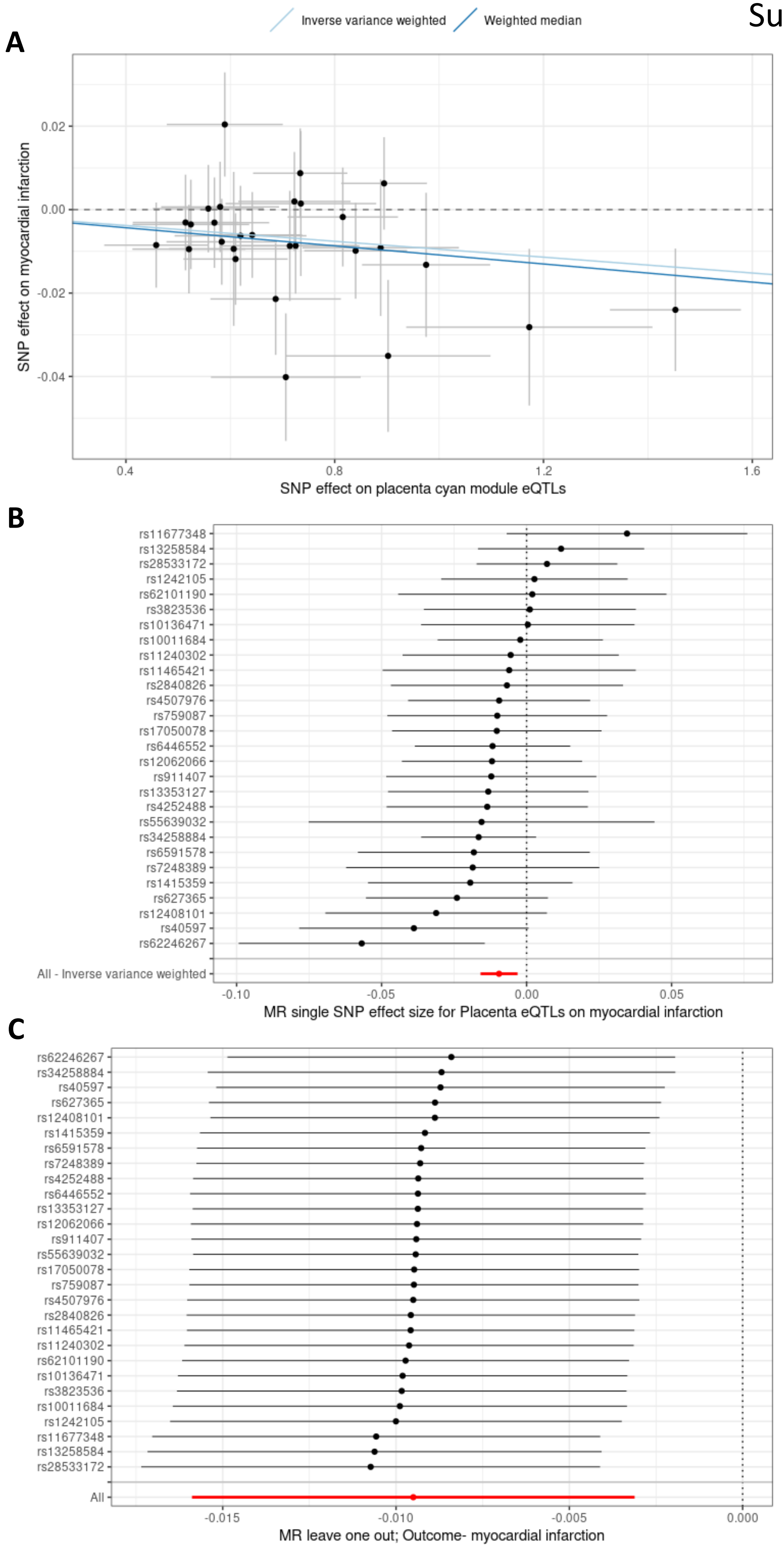

Supp Figure 6

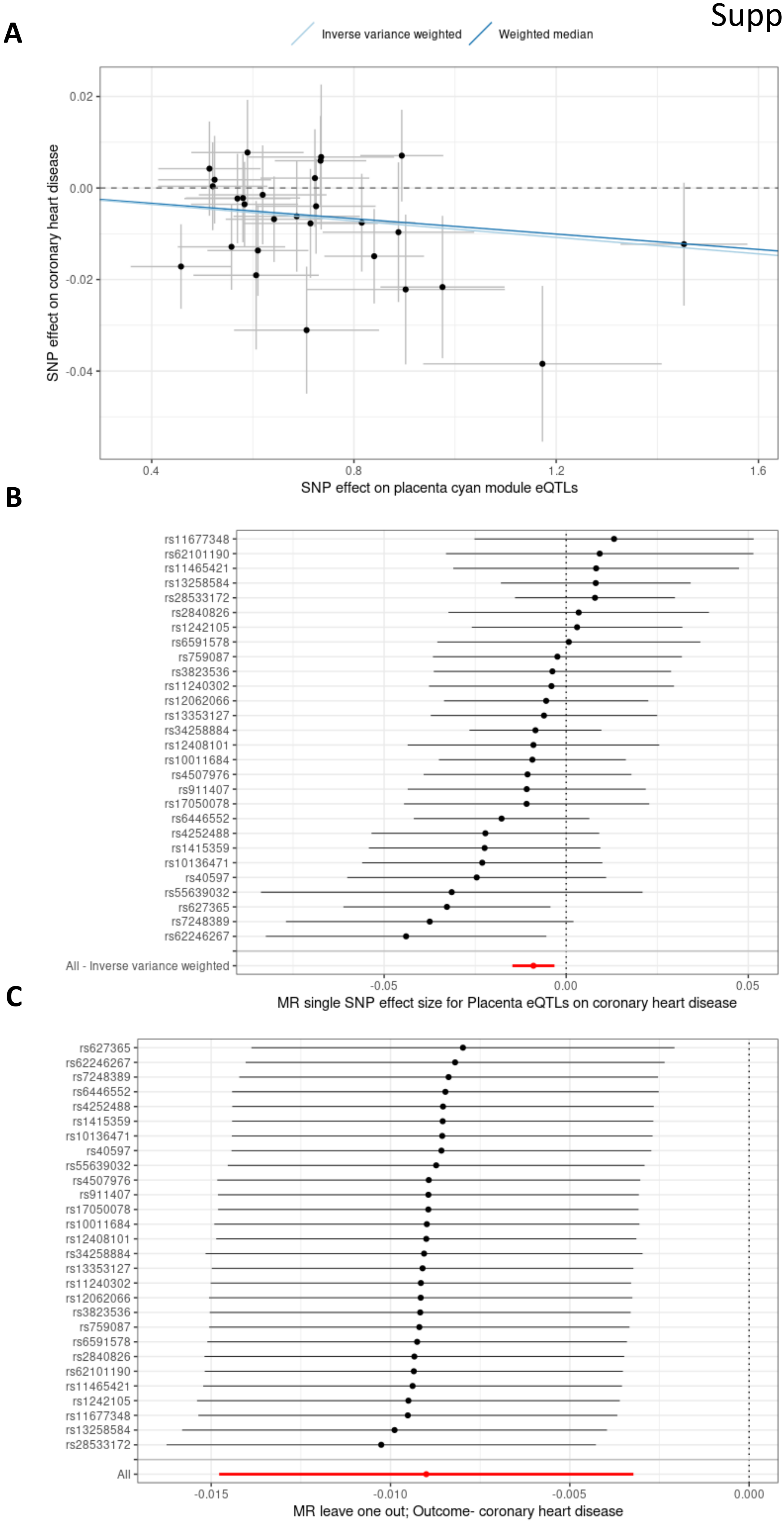

Supp Figure 7

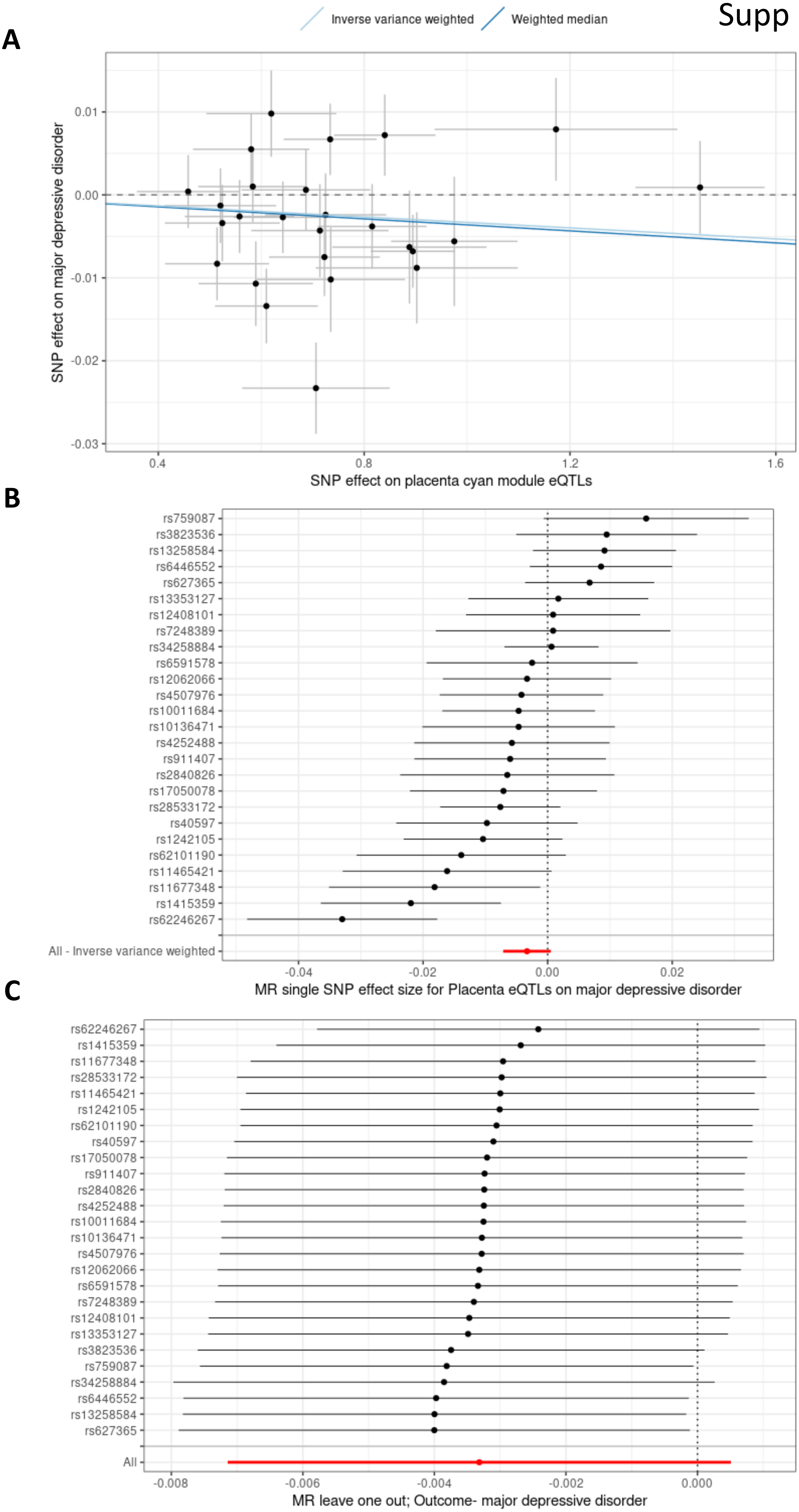

Supp Figure 8

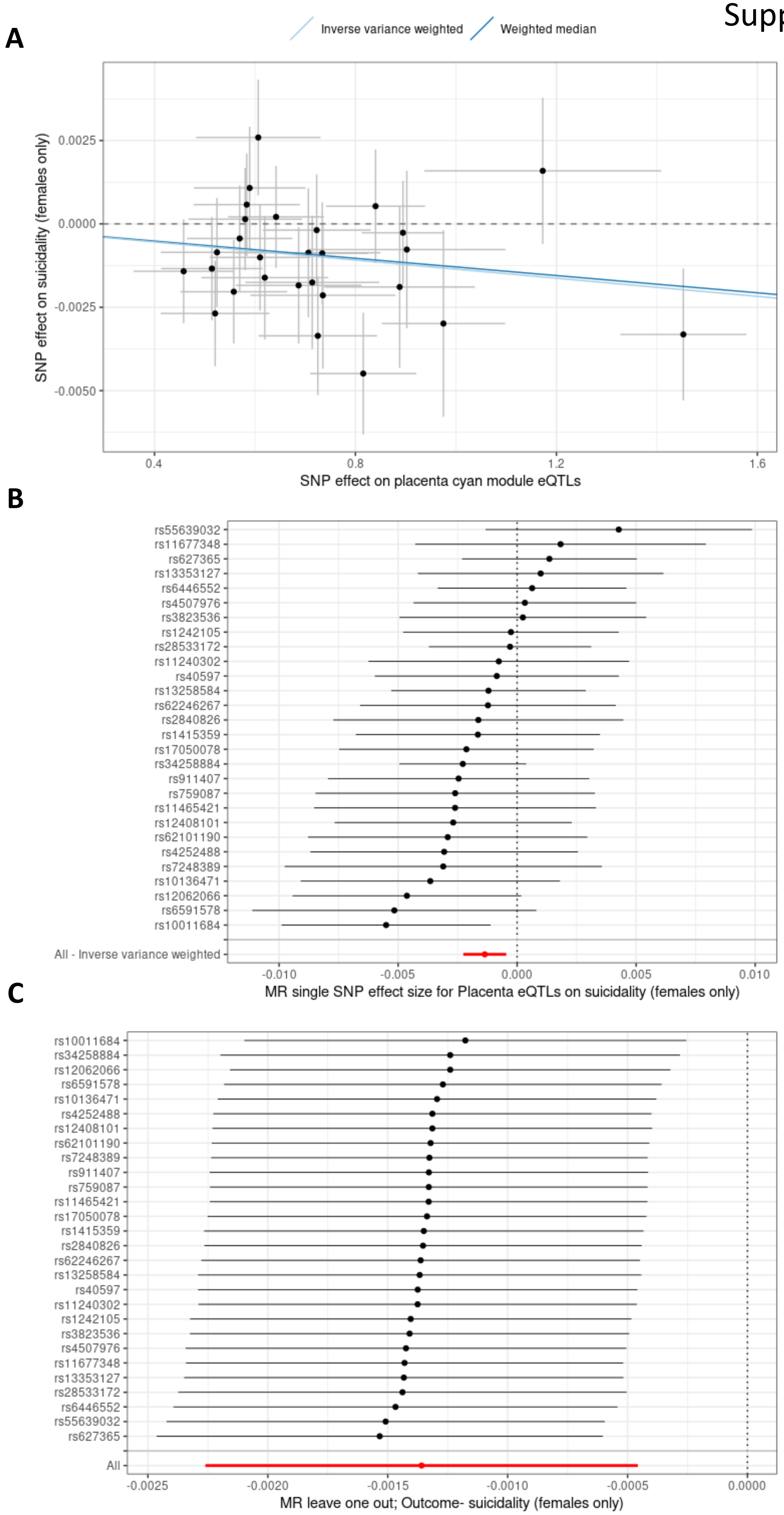

Supp Figure 9

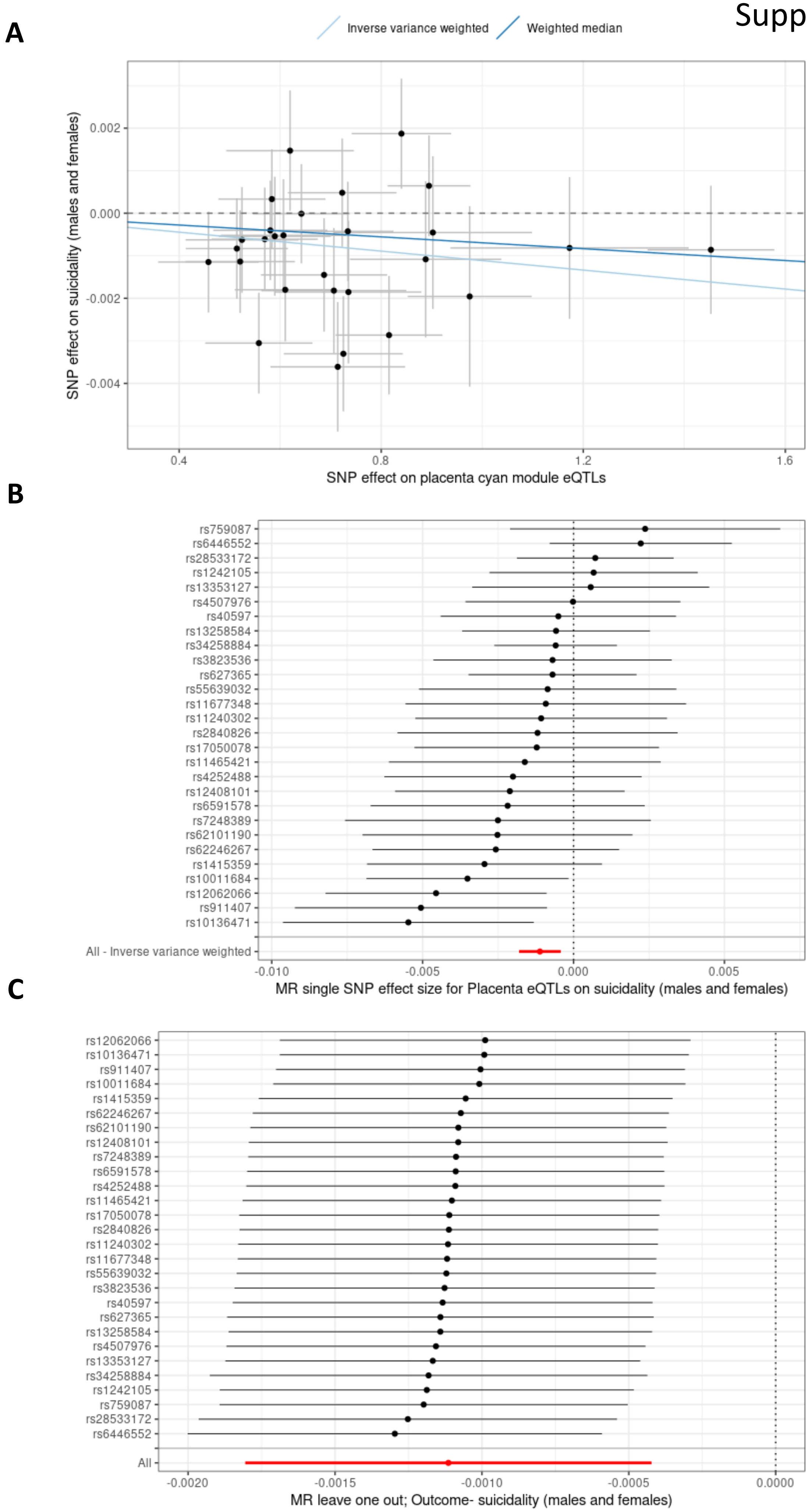
